# Impact of the Pandemic: Screening for Social Risk Factors in the Intensive Care Unit

**DOI:** 10.1101/2022.06.01.22275889

**Authors:** Derek Ge, Alec M. Weber, Jayanth Vatson, Tracy Andrews, Natalia Levytska, Carol Shu, Sabiha Hussain

**Affiliations:** Department of Medicine, Robert Wood Johnson Medical School, Rutgers University, New Brunswick, NJ 08901, USA; Department of Medicine, Westchester Medical Center, Valhalla, NY 10595, USA; Department of Biostatistics and Epidemiology, School of Public Health, Rutgers University, Piscataway, NJ 08854, USA

## Abstract

Due to limitations in data collected through electronic health records, the social risk factors (SRF) that predate severe illness and restrict access to critical care services are poorly understood. This study explored the feasibility and utility of directly eliciting SRF in the ICU by implementing a screening program. 566 critically ill patients at the medical ICU of Robert Wood Johnson University Hospital from July 1, 2019, to September 31, 2021, were screened for seven SRF. We compared characteristics between those with and without each SRF through Chi-squared tests and Wilcoxon Rank Sum tests. Overall, 39.58% of critically ill patients reported at least one SRF. Age, socioeconomic status, insurance type, and severity score differed significantly depending on the SRF. Most notably, the prevalence of SRF, overall and individually, changed after March 2020 which represented the onset of the COVID-19 pandemic. Our findings indicate that SRF can induce low-risk severe illnesses and restrict access to critical care services.

## INTRODUCTION

Social determinants of health (SDOH) are defined by the World Health Organization as “the conditions in which people are born, grow, live, work, and age.”^1^ When SDOH are responsible for premature morbidity and mortality, they are referred to as social risk factors (SRF).^2^ Modifiable SDOH – subject to culture, policy, and institutions – have been found to outperform measures of access and quality of clinical care in predicting longevity and well-being.^3^ However, a minority of hospitals and physician practices alike identify and address SRF.^4^ The reluctance to consider SDOH beyond basic sociodemographic data (e.g., age, race, and gender) and reliance on physician notes to record SRF reflects a lack of standardized assessment tools and the limited scope of modern electronic health records (EHR).^5-7^

The intensive care unit (ICU) often admits patients with critical illness potentiated by SRF, most commonly due to sepsis, but the extent to which critical illness can be attributed to SRF has not yet been satisfactorily demonstrated.^8,9^ Since the burden of SRF falls inequitable upon blacks in the United States, many studies have explored whether racial disparities exist in the ICU.^10^ However, associations between race and mortality were strongly attenuated by socioeconomic status (SES), insinuating that race alone cannot fully capture the complex relationship between SDOH and health.^11^ While SES may more accurately reflect the distribution of SRF, most literature in critical care medicine represent SES using abstracted indexes from census data.^12-16^ Problematically, this assumes that all residents of a neighborhood share the same SRF and reinforces the stereotype that only low-income families experience SRF.^17^

Rather than approximating SRF from basic sociodemographic data or SES, we implemented a screening program to directly discuss multiple domains of SDOH with our patients. The pertinence of our goal became more apparent in the wake of the social impact inflicted by the COVID-19 pandemic. Economic hardships, often brought by loss of employment, left families struggling to pay for rent and food.^18^ Certain communities, in color or creed, were disproportionally affected by the virus from reasons including the inability to enforce physical distancing and a greater burden of comorbidities.^19^ Patients in these communities were more likely to become hospitalized because of COVID-19 and later admitted to the ICU or die.^20,21^ Given the role the ICU serves in treating the most severe cases of community-acquired illnesses, changes in the proportions of patients with SRF could reflect reverberations from catastrophes and policies at the national and local level.

Importantly, we sought to determine the feasibility and utility of eliciting SRF from critically ill patients. The primary objectives of this single center prospective observational cohort study were to 1) quantify the prevalence of SRF in the ICU, 2) determine what characteristics are associated with SRF, and 3) contextualize our findings regarding policy responses to COVID-19.

## METHODS

### Study Design and Participants

From July 1, 2019, to September 30, 2021, all patients admitted to the medical ICU at Robert Wood Johnson University Hospital were eligible for this prospective observational cohort study. Surgical patients are monitored in a separate department and were not considered for our study. For incapacitated patients, our team interviewed visiting family members. We excluded patients who were without family or refused participation. Because our study intended to connect New Jersey (NJ) residents to local resources, we only included patients with addresses within NJ. Finally, screening was only conducted when a social worker was available, excluding patients who were only available for screening at night or during the weekend.

Our study includes patients treated prior to and during the pandemic. We marked the onset of the pandemic as March 30, 2020, when patients were first admitted to the ICU with COVID-19 related illness. Due to the influx of these admissions in the beginning of the pandemic, we temporarily halted data collection from March 30, 2020, to June 30, 2020.

Participants were asked questions based off an adapted version of the American Academy of Family Physician’s Social Needs Screening Tool borrowed from our healthcare system’s family-oriented community clinic, the Eric B. Chandler Health Center (Exhibit 1). This comprehensive screening tool was selected to capitalize on existing partnerships with community organizations that offered support for disadvantaged patients. Multiple domains of SDOH were covered by the screening tool: food, housing, utilities, medication, transportation, access to healthcare, and caregiver support.

**Exhibit 1.**
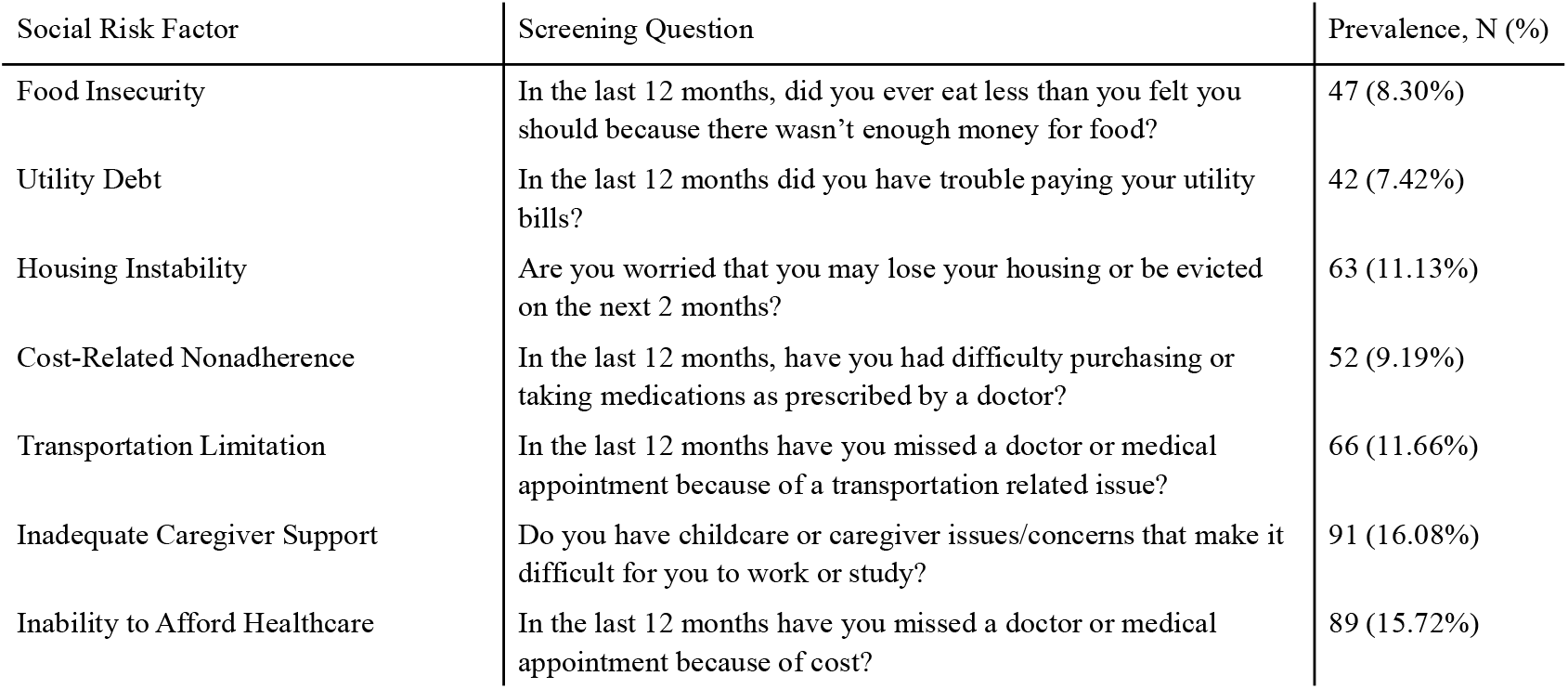
Screening Tool for Social Risk Factors

To successfully incorporate screening into the ICU workflow, we contacted IT staff and administration for support in embedding the screening tool into our EHR. Prior to engagement, we held teaching sessions to promote teamwork and reduce communication errors between residents, nurses, pharmacists, social workers, case managers, and students. Following a script, an attending physician, critical care fellow, or nurse interviewed each patient and provided the opportunity to consult with social services. For patients who did not prefer to use English during the clinical interaction, we consulted interpreters through LanguageLine Solutions.

### Data

Sociodemographic data (age, race, gender, and insurance type) were extracted from electronic medical records. In the EMR, racial identity was often assumed rather than solicited, limiting our ability to make detailed comparisons across race and ethnicity. Unfortunately, this is a pervasive aspect of studies like ours that rely on staff observations instead of self-report for determining racial identity.^22^ As such, we used a dichotomous race measure, denoting if a person was Black or African American. Public data from the U.S. Census Bureau’s American Community Study was used to match zip codes with median income per household (ZMHI). ZMHI was divided into four quartiles with Q1 representing the poorest areas serviced by our healthcare system. To describe the clinical presentation of each patient, we used the Acute Physiology and Chronic Health Evaluation III (APACHE III) scoring system for its superior discriminatory ability.^23,24^ Dichotomous variables were created to indicate which SRFs were endorsed by individuals.

### Statistical Analysis

Univariate statistics including frequency (%) and median (IQR) were calculated for all variables; normality was assessed using Shapiro-Wilks test (p < 0.05). Comparisons across SRF groups were performed using Chi-squared tests for categorical data and Wilcoxon Rank Sum tests for continuous data. Pearson correlation matrices were used to evaluate relationships among SRF groups and personal characteristics. To determine shifts in the prevalence of SRF prior to and during the pandemic, we compared the proportion of patients in each period who screened positively for SRF. All analyses were performed in R Studio (4.0.3) and used two-tailed tests with an alpha of 0.05. To note, our results differ from an earlier analysis published in an abstract due to refinement of our exclusion criteria and timepoint definition.^25^

## RESULTS

### Study Population Characteristics

Out of 1319 ICU admissions, 566 (42.91%) NJ residents underwent screening during our study period. Most of those screened arrived after the onset of the pandemic (443, 78.27%) and a sizable minority reported at least one SRF (224, 39.58%). Exhibit 1 displays the seven SRF elicited by our screening tool along with the corresponding questions. The percentage of patients attributed to each SRF was lowest for utility debt (42, 7.42%) and highest for inadequate caregiver support (91, 16.08%).

In Exhibit 2, we highlight demographic and clinical differences between patients with and without SRF. Patients who reported SRF were, on average, younger except for those with the inability to afford healthcare, who were significantly older (Exhibit 2A). Except for patients reporting inadequate caregiver support, those with SRF were disproportionally represented in the poorest two ZMHI quartiles (Exhibit 2B) and were more inclined to use Medicaid or out-of-pocket sources of healthcare coverage (Exhibit 2C). The median severity score was lower for patients with SRF compared to those without (Exhibit 2D). Gender did not significantly differ between patients with or without SRF. The proportion of black patients only diverged in one domain of SDOH, healthcare access, in which it was considerably lower (5.62% vs. 15.51%). Conversely, the percentage of white patients with the inability to afford healthcare was disproportionally high (70.79% vs. 61.01%).

**Exhibit 2.**
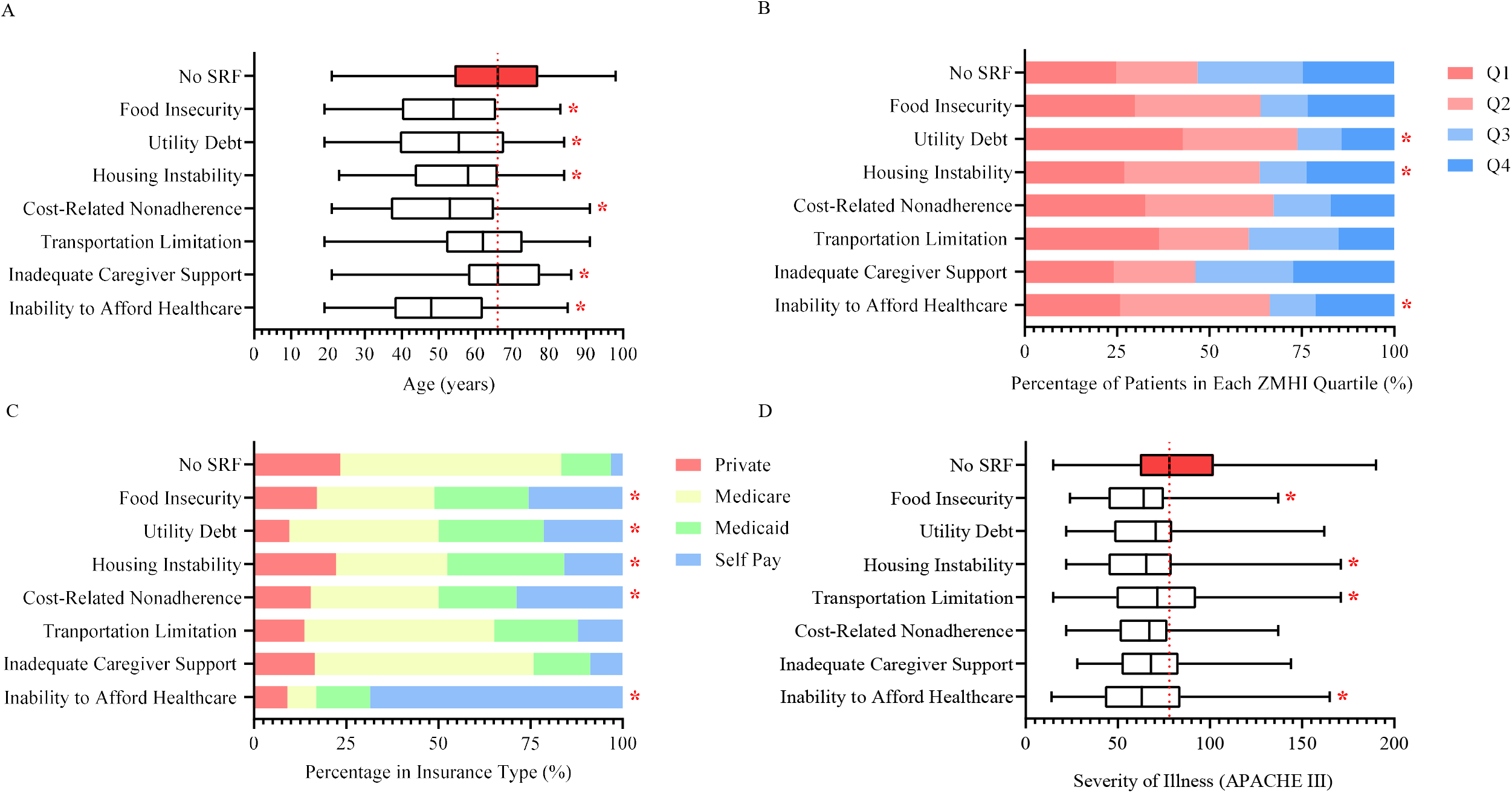
Characteristics of Patients with Social Risk Factors

### Correlation Matrix

SRF were weakly to moderately intercorrelated (Exhibit 3). Food insecurity, utility debt, housing instability, cost-related nonadherence, and transportation limitation were positively interrelated with strengths ranging from 0.27 to 0.57. Inability to afford healthcare was not related to three other SRF and inadequate caregiver support was weakly correlated with other SRF except for transportation limitation (r < 0.20). Patients with SRF trended towards reporting either inadequate caregiver support, inability to afford healthcare, or a combination of the remaining SRF targeted by our screening program.

**Exhibit 3.**
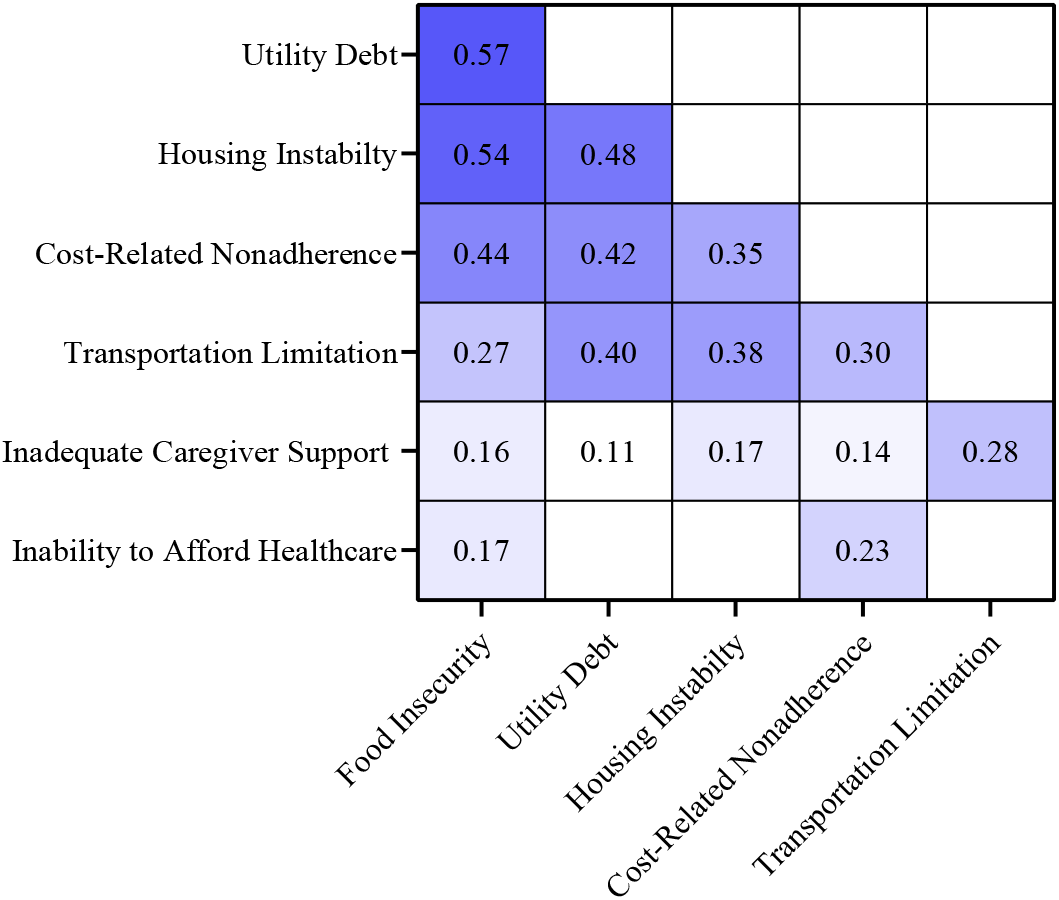
Correlation Matrix of Social Risk Factors

### The Impact of the Pandemic

The social impact of the COVID-19 pandemic influenced access or demand for critical care services, as shown in Exhibit 4. The prevalence of SRF in our ICU fell from 54.47% to overall 35.44% and decreased across all categories except for inability to afford healthcare, which increased from 7.32% to 18.06%.

**Exhibit 4.**
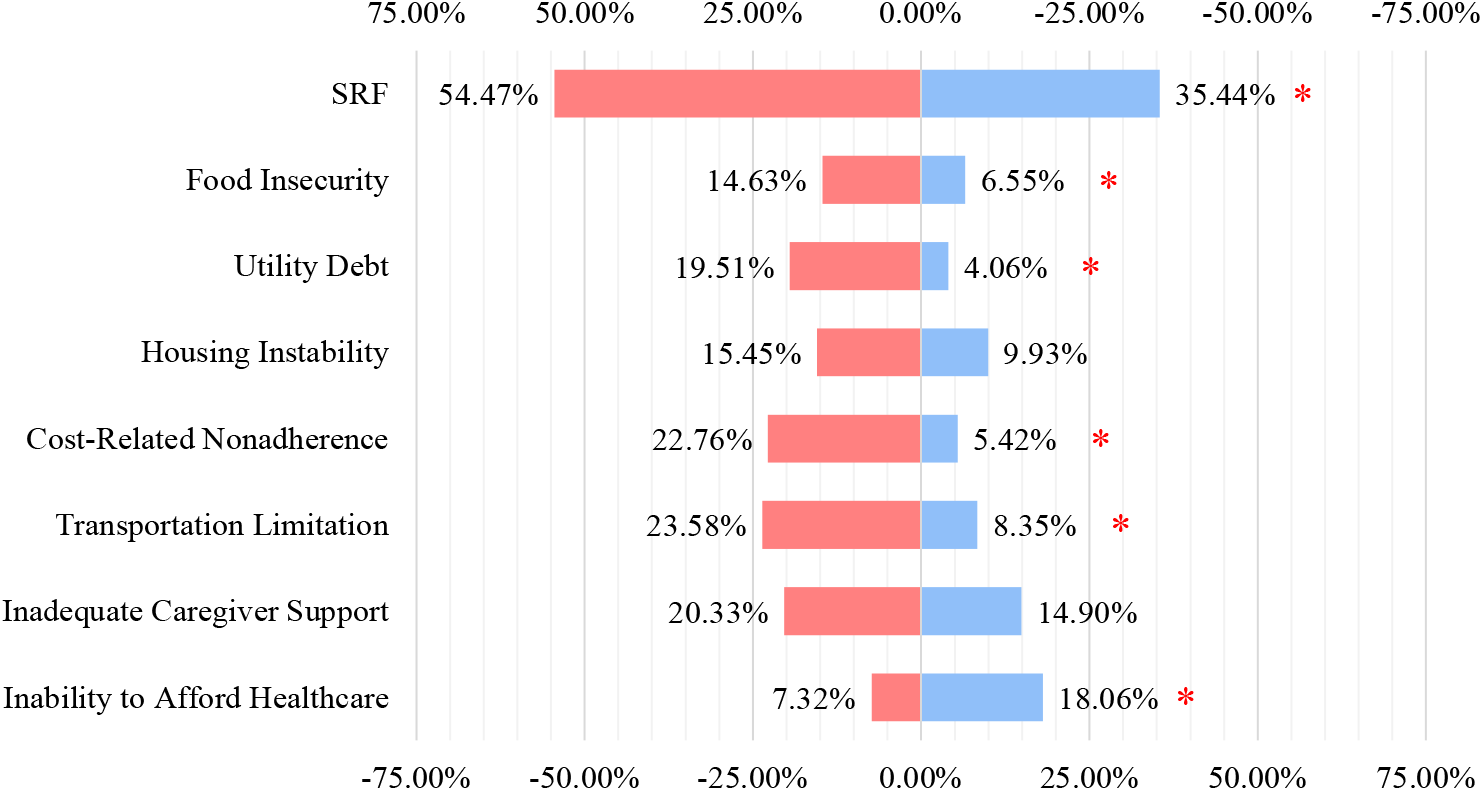
Prevalence of Social Risk Factors Before and During the Pandemic

## DISCUSSION

Relying on primary care services for screening assumes access to healthcare, which alienates those who may benefit the most from being connected to social services. Discussing SDOH with critically ill patients and their families defined norms on both sides of the clinical interaction. As such, our study required months of preparation to embed a screening tool into our EHR and educate all members of our clinical team. However, by contextualizing ICU admissions with SRF, we enabled our healthcare system and community partners to respond accordingly. Our study is novel for 1) introducing a screening program for multiple SRF into the ICU and 2) exploring the characteristics and prevalence of critically ill patients with SRF during an ongoing pandemic.

The fallout from the COVID-19 pandemic demonstrates how the ICU can function as a barometer of social change. From March 2020 to April 2020, NJ witnessed a record high unemployment rate and unprecedented numbers of unemployment insurance claims, leading to large losses in employer-based health coverage.^26^ During the same time, the State Department of Health, under the leadership of Governor Philip Murphy, released triage guidelines prioritizing younger patients with lower severity scores.^27^ These conditions may have produced an overwhelming amount of newly uninsured patients who were prioritized for ICU admission given their favorable prognosis, producing a shift in our patient population. Meanwhile, the overall prevalence of SRF reported by critically ill patients fell by about 20% since the pandemic. Optimistically, this may reflect the work of organizations, energized by investments from the New Jersey Pandemic Relief Fund and American Rescue Plan, that launched programs for food distribution, legal aid enforcing rent moratoriums, and assistance with affording medications.^28,29^ Conversely, this trend could indicate exacerbated inequity in healthcare access. During shortages in ICU bed supply, Kanter et al. identified a large gap in access by income, despite COVID-19 disparately harming low-income households who were more likely to report SRF.^30,31^ By studying SRF in an ICU setting, we continue the narrative that allocation criteria failed to prioritize distributing vaccines and critical care services to disadvantaged individuals and groups.^32^

Notably, critically ill patients with SRF in our cohort were disproportionally assigned a low severity score upon admission. While the dearth of studies on SRF in the ICU prevents a conclusive explanation, this finding may be an artifact of a distinct etiology. Perhaps the youth of patients with SRF, along with differences in their profile of comorbidities, diminished the odds of mortality.^33,34^ Interestingly, Bein et al. concluded that critically ill patients with a low SES, as determined by a questionnaire, had increased risk of a high severity score.^35^ We attribute our contradictory findings to our decision to conduct our study in the medical ICU, which receives patients with community-acquired disease rather than surgical complications. Furthermore, we directly elicited SRF instead of making assumptions by SES. Although healthier patients require fewer diagnostic tests and equipment, these patients still accumulate high levels of fixed overhead costs that constitute the bulk of ICU spending.^36,37^ Critical care medicine accounts for 13.21% of total hospital care expenditure as costs per day in the ICU equals six to seven times those in a general ward.^38,39^ Many institutions have concluded that treating low-risk patients in the ICU is not cost-effective, instead opting for an intermediate or step-down unit.^40,41^ Thus for healthcare systems, there exists a financial imperative to address SRF that discourages proper health management, ultimately leading to possibly preventable ICU admissions.

In an era of precision medicine and growing recognition of SDOH, the contemporary clinical workflow in the ICU should include a standardized process to detect SRF. Our study demonstrates that screening for SRF vastly augments the relevance of basic sociodemographic data and SES. For example, inadequate caregiver support may predominantly affect elderly Medicare beneficiaries with multiple chronic conditions that can be exacerbated without proper monitoring and support.^42^ Patients with this SRF were older and not defined by a low SES. On the other hand, those with the inability to afford healthcare were younger and disproportionally represented in the lowest two ZMHI quartiles. We found that most patients with the inability to afford healthcare were white; however, we hypothesize this number could include many Hispanics, who are disproportionally uninsured or disqualified due to immigration status.^43,44^ We felt comfortable in this assumption given that the largest ethnic group within our zip code are Hispanics.^45^ That SRF could represent the unmet needs of divergent subgroups in a heterogenous critically ill patient population led us to explore intercorrelations between SRF. Surprisingly, we found that certain SRF – but not all – shared modest correlations and were often reported together. This implies that policy addressing SRF should carefully consider the domains of SDOH relevant to the target population.^46^

## Limitations

As a single-center investigation, immediate generalization of the presented findings to other institutions has not yet been validated. However, we share the results of our study to encourage other ICUs to identify SRF in their patient population. Further, our definition of racial and ethnic identity is limited due to incomplete data collection, especially because our failure to qualify Hispanic versus non-Hispanic. However, this reflects oversight by our healthcare system and others.^47^ That race and ethnicity are assumed by the admitting clerk in most hospitals calls into question the validity of conclusions based on EHR data on race alone.^22^

## Conclusion

Neglecting to acknowledge SDOH in the ICU may perpetuate ignorance of SRF that predispose disadvantaged patients towards developing low-risk severe illnesses, incurring high medical costs, and requiring critical care services. In our prospective observational cohort study, we successfully introduced a standardized procedure for discussing SDOH in our ICU workflow by implementing a screening program at our medical ICU endorsed by administration, IT staff, and our clinical team. Importantly, we found that while nearly four in ten critically ill patients reported at least one SRF, the overall prevalence of SRF changed from structural forces set in motion by the COVID-19 pandemic.

## Data Availability

All data produced in the present study are available upon reasonable request to the authors.

## Notes

### Competing Interest Statement

The authors have declared no competing interest.

### Funding Statement

This study did not receive any funding.

### Author Declarations

Ethics committee/IRB of Rutgers Biomedical and Health Sciences gave ethical approval for this work.

